# Charting Champions: Online Coaching to Reduce Physician Administrative Burden and Improve Well-Being

**DOI:** 10.64898/2026.08.05.26359826

**Authors:** Sarah Smith, Daniel Lemoine

## Abstract

**Objective:** To assess the efficacy of an executive peer coaching program, Charting Champions Program (CCP), in helping physicians manage their administrative workload, thereby improving time management, workflow and well-being.

**Findings:** In this longitudinal survey study, physicians self-reported significant improvements in completing charting and administrative paperwork during their clinical day. Physicians reported significant improvements in mental, cognitive and emotional states after the program.

**Meaning:** The Charting Champions Program is an effective intervention that supports physicians in problem-solving the administrative burden of their clinical day, improving workflow efficiency, completing administrative requirements during clinical hours, and enhancing work-life balance and personal satisfaction.

**Background:** Physicians are subject to high levels of mental, physical, and emotional stress, partly due to increasing administrative burdens. Online coaching is a proven intervention to help physicians improve workflow efficiency, reduce administrative burden and improve job satisfaction.

**Design:** This voluntary longitudinal survey took place between 2020 and 2023. Physicians were asked to complete a survey at program entry and again 30-90 days after program completion. The survey consisted of 14 Likert scale questions, and a final sample of 280 physicians completed both surveys.

**Intervention:** CCP contains modules that teach workflow improvements for clinical days, including timely charting, administrative task workflow, managing patient consultations and reducing interruptions. Interventions include self-paced modules, live coaching, recordings and an online peer community.

**Results:** Post-CCP physicians reported a significant decrease in hours spent charting (P<0.0001) and completing clinical paperwork outside of clinical hours (P<0.006). Physicians also reported a decrease in work-related dread (P<0.001), feelings of burnout (P<0.001), and thoughts of quitting due to administrative burdens (P<0.001). Physicians felt more focused at work (P<0.001), felt more in control of the clinical day (P<0.001), and rated their mental energy at work higher (P<0.001). The program did not affect the number of patients seen in a full clinical day (P > 0.918).

**Conclusion and Relevance:** The CCP reduces the time physicians spend on tasks outside of clinical hours, increasing free time without decreasing the number of patients seen per day.

## Introduction

Physicians play an indispensable role in society by providing essential healthcare services, diagnosing and treating illnesses, and promoting overall well-being. However, physicians often contend with heavy workloads, long hours, and the emotional toll of patient care. High administrative burdens, inadequate work-life balance, and insufficient mental health support further complicate their professional lives. The sustainability of healthcare systems relies on finding elegant solutions to help physicians cope with the evolving demands of their profession. ^1–4^

Excessive administrative burdens, including time spent on electronic health records (EHRs) and other clerical tasks, are detrimental to physician well-being. Previous research has demonstrated that physicians spend more than half of their work time on EHR-related tasks^4–8^. Physicians report that managing EHRs can turn them into “unhappy data-entry clerks”^24^ rather than allowing them to focus on clinical responsibilities. The burden of EHR clerical tasks is also linked to decreased job satisfaction and professional fulfilment, and lower satisfaction with their work-life integration, including greater feelings of ineffectiveness ^2,9^.

Peer coaching, which includes Clinical Day Advice and Mentorship, can help physicians manage these administrative burdens by enhancing their time management skills, increasing their efficiency, and fostering a greater sense of control over their work environment ^23^. Coaching provides physicians with strategies to prioritise tasks, delegate non-essential administrative work, and streamline their workflows ^25^. Prioritisation strategies and delegation skills not only reduce the time spent on paperwork and other clerical duties but also alleviate the stress associated with these tasks. Additionally, coaching helps physicians develop resilience and more effective coping mechanisms, allowing them to handle the pressures of their administrative responsibilities more effectively ^10–15^.

The Charting Champions Program (CCP) is an exclusively online coaching program that begins with a foundation of common knowledge base skills in clinical day problem-solving for its physicians. The CCP foundational modules help create frameworks for workflow improvements in charting, time management, scheduling, administrative task management, reducing interruptions, and managing incomplete tasks. Physicians in the CCP program also become part of an active online community of colleagues supported by the program’s coaches and clinical day advisors. The CCP setting fosters self-awareness, helping individuals to identify their strengths and areas for improvement. Through coaching, physicians set clear, achievable goals and develop actionable plans to reach them.

## Methods

### Physicians

We invited N=1297 clinical physicians from Canada and the US who were between 1 and 15 years post-residency and had participated in the CCP program from 2020 to 2023 to complete a two-part voluntary pre-post online survey. The survey responses were anonymous and were initially collected for program evaluation and improvement.

### Study Design

We completed a longitudinal analysis on a survey distributed to physicians. The surveys asked physicians to outline their current experience of their clinical day, before the intervention and 30-90 days after completion of the foundational core modules. Only the results from the 280 physicians who completed both the pre-CCP and post-CCP surveys were analysed for this study.

### Intervention

The online Charting Champions Program contains five foundational modules that provide specific frameworks for problem-solving on the clinical day and for assisting physicians in improving workflow. Modules teach physicians how to: (1) complete their charting immediately following patient interactions; (2) evaluate and lead their consultations to run closer to time; (3) manage the administrative tasks of the day more efficiently; (4) develop strategies for reducing interruptions; and (5) develop a process for managing incomplete tasks that have accumulated. In addition, the program provides group coaching through six to eight calls each month, facilitated by the physician coaches. These calls can be accessed live, and the recordings can be reviewed later. If physicians desire additional coaching, they can utilise a program-specific online community to access peer support and written coaching.

### Main Outcomes and Measures

The primary outcomes were self-reported changes in the number of hours spent charting and doing paperwork outside clinical hours, as well as changes in feelings of burnout, quality of life, work-related dread, sense of control, and focus. Secondary outcomes included changes in charting experiences, inbox states, resignation ideation, and quantity of physicians’ free time.

### Data Analysis

The survey consisted of 14 Likert-scaled questions. We compared the responses on the Pre and Post surveys (prior to and 30-90 days after completion of the core modules) using either T-tests or Chi-squared tests. Questions asking physicians for ordinal numerical responses (e.g. How many hours per week do you spend charting outside clinical hours?) were converted to interval data and analysed using the student’s t-test.

Alternatively, questions asking physicians to rank their responses on a purely ordinal scale (Never/Rarely/Sometimes/Frequently/Always) were analysed using Chi-squared tests, comparing physicians who answered Never/Rarely to those who answered Frequently/Always (neutral responses removed).

## Results

1,297 physicians were invited to complete the pre/post-coaching Charting Champions surveys; 980 completed the pre-intervention survey, and 296 completed the post-intervention survey. 280 physicians completed both surveys and were included for analysis in this study (**Figure 1**). Physicians who completed both studies reported a reduction in hours spent charting outside clinical hours (−0.42 h; 95% CI, −0.58 to −0.25 h; P < 0.001; **Figure 1**). Post-CCP physicians also reported fewer hours spent doing paperwork outside clinical hours (−0.23 h, 95% CI: −0.40 h to −0.07 h, P = 0.006, **Figure 1**). Physicians reported no significant difference in the number of patients seen in a normal full clinical day (−0.10, 95% CI: −0.17 to +0.16, P = 0.918, **Figure 1**).

**Figure 1.**
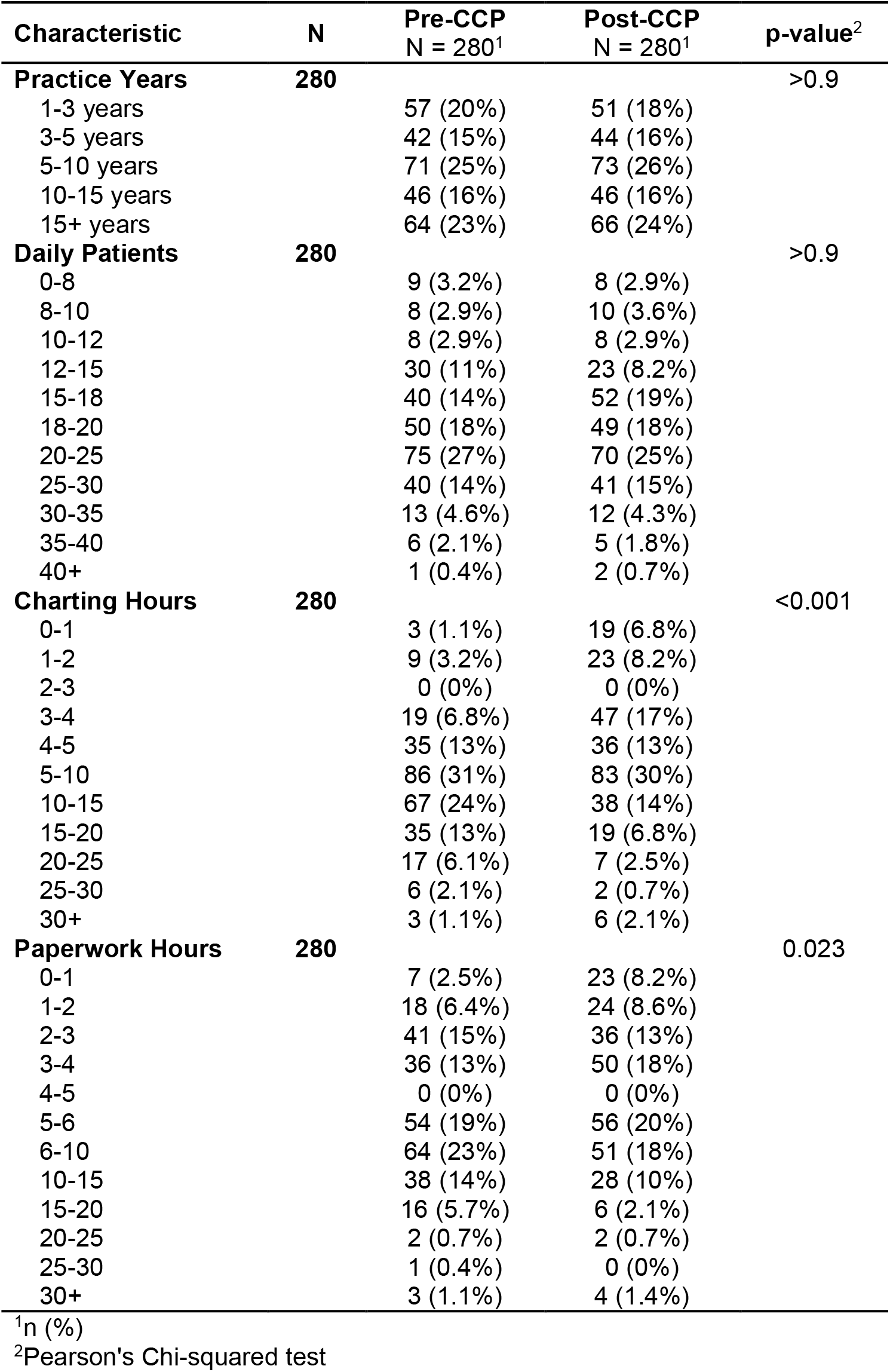
Survey Responder Characteristics, before and after participating in the Charting Champions Program. Characteristics of physicians at the time of Pre-CCP and Post-CCP surveys. The table only includes results from physicians who completed both surveys.

After completing the Charting Champions program, physicians were less likely to dread going to work (p<0.001, **Figure 2**) or to frequently consider quitting their jobs (p<0.001, **Figure 2**). Physicians also self-reported being less frequently burnt out (p<0.001, **Figure 2**). The proportion of physicians who felt focused at work increased (p<0.001, **Figure 2**) along with the proportion of physicians who felt in control of their clinical days (p<0.001, **Figure 2**). The proportion of physicians who frequently or always had their evenings/weekends free of paperwork increased from 16% to 30 (p<0.001, **Figure 2**). Post-intervention, physicians were also more likely to report their mental energy at work as high or very high (p<0.001, **Figure 3**).

**Figure 2.**
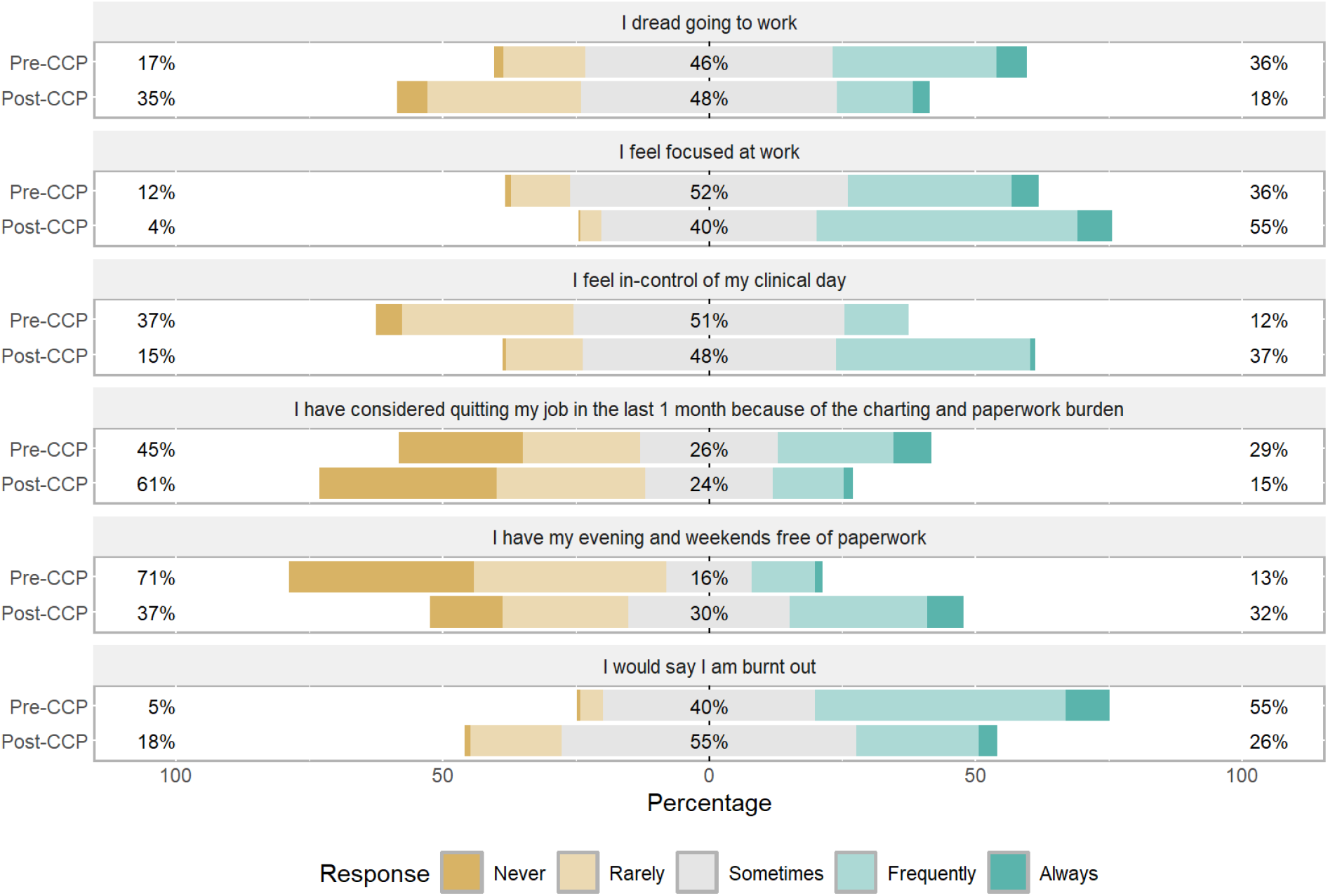
Survey responses. Proportion of physician responses (N=280) to ordinal scale survey questions, percentages indicate proportion of physicians who answered Never/Rarely, Sometimes, or Frequently/Always.

**Figure 3.**
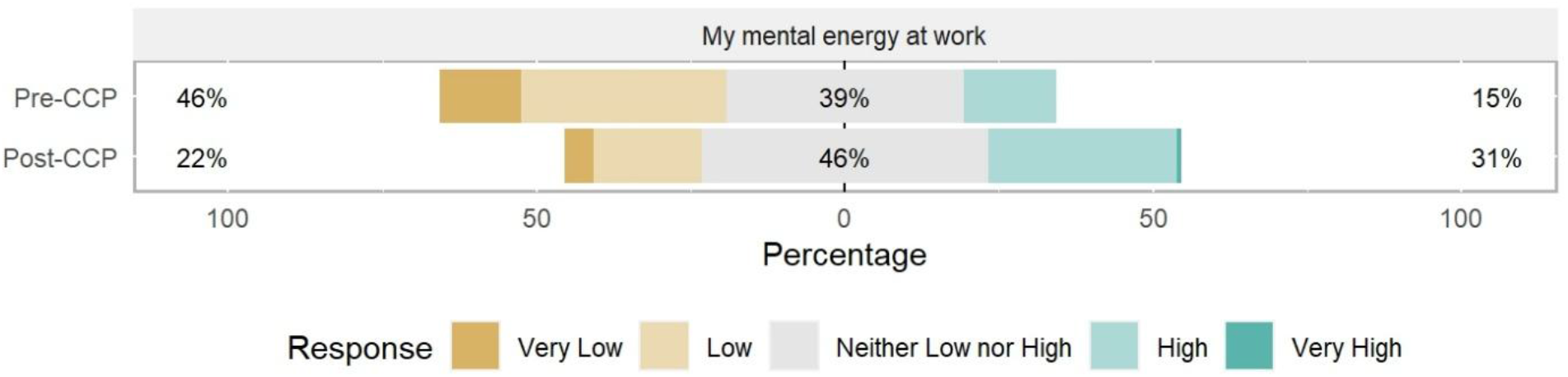
Mental Energy at Work. Proportion of physician responses (N=280) when asked to rate their mental energy at work, percentages indicate proportion of physicians who answered Very Low/Low, Neither Low nor High, or High/Very High.

## Discussion

Data collected from Charting Champion Program member evaluations supports the growing evidence in the literature that coaching can significantly improve the well-being of physicians ^10, 13–15,22–29.^.Upon trialling and implementing the strategies designed with their coaches, physicians reported an improved ability to manage their daily administrative tasks (e.g., charting, in-basket, and emails) and to address incomplete tasks that accumulate over time. The CCP results demonstrate that physicians reported increased personal time. The CCP intervention was effective in reducing feelings of dread, burnout, and thoughts of resigning, while also improving motivation, control, and mental energy at work.

Physicians in Canada and the US are becoming overwhelmed by rising patient workloads ^1–3,20^ while navigating a complex and evolving array of operational requirements. These requirements include more patient interactions, handling governmental and insurance billing systems, keeping up with changing record-keeping practices, and managing limited resources and inadequate support structures. As a result, physicians are experiencing significant levels of mental, physical, and emotional stress ^2^. Many organisations and researchers are striving to understand the impact of these challenges on physicians ^19,21^ and are developing and testing various potential solutions, including peer coaching.

The Charting Champions physician peer coaching program is an innovative online initiative designed to help physicians manage the demands of their clinical day, reduce time spent working outside clinical hours, and improve overall well-being in their current work environment. Physician coaching (targeting administrative and/or personal improvements) is a proven intervention that positively impacts patient care and physician work-life balance ^6,15–19^. We found that the CCP was effective in reducing the number of hours physicians spent on administrative activities outside clinical hours. The program also improved physicians’ outlook and didn’t reduce the number of patients they could see per day.

Coaching programs like the CCP create personalised, supportive educational environments that help physicians identify and address inefficiencies in their clinical day through a peer coach. Coaching programs are designed to leverage physicians’ natural mental and emotional strengths, as well as their prior life and work experiences, to problem-solve and develop strategies for better balancing their organisational and professional demands.

### Limitations

This study was limited to voluntary responses to multiple surveys from physicians who participated in the CCP program, resulting in a self-selected sample. To ensure anonymity in the dataset, factors such as gender, specialty, and practice location were not collected. The generalizability of our findings to specific groups could be limited.

## Conclusion

While physician coaching occurs on an individual basis, it can improve the functioning of healthcare organisations by enhancing physician retention and productivity. Coaching helps improve physicians’ ability to adapt to changing administrative requirements and to sustain large clinical workloads. We demonstrate that the Charting Champions Program is an effective intervention to improve physician outcomes in modern clinical practice without adversely affecting their productivity.

## Acknowledgements

The authors would like to extend thanks to the physician coaches and support staff who participated in delivering this coaching program. A special thanks to Harrison Smith BSc, MBioMedSci, for editorial assistance.

## Data availability

The datasets analysed for this study are available from the corresponding author on reasonable request.

## Declarations

### Ethics Approval

This project utilized de-identified datasets provided by a private corporation under pre-existing participant consent. Because the study relied entirely on anonymized data and collected no new sensitive information, it carried negligible risk. Consequently, the project was deemed exempt from the Human Research Ethics Committee (HREC) review, in accordance with the NHMRC National Statement on Ethical Conduct in Human Research.

